# Common Variant Contributions to Neurodevelopmental Risk in Orofacial Clefts

**DOI:** 10.64898/2026.05.05.26352467

**Authors:** Alexandros Rammos, Sarah J Lewis, Amy Davies, Yvonne Wren, Kerry Humphries, Jonathan Sandy, Gemma C Sharp, Michael J Owen, Marianne B M van den Bree, Evie Stergiakouli

## Abstract

**Background:** Children with cleft lip and/or palate (CL/P) experience increased rates of neurodevelopmental difficulties, including ADHD, autism spectrum disorder, and educational challenges. While rare neurodevelopmental copy number variants (ND-CNVs) are enriched in this population and associated with poorer outcomes, these variants are present in only a small proportion of children born with cleft. Whether shared common genetic variation contributes to neurodevelopmental comorbidities in CL/P remains unknown.

**Methods:** We investigated this question using data from 2,313 children with CL/P from the Cleft Collective and 7,913 population controls from the Millennium Cohort Study. We tested for shared genetic architecture using linkage disequilibrium score regression, examined associations between polygenic risk scores for eight cognitive, neurodevelopmental, and psychiatric traits and developmental and behavioural outcomes within the cleft population, compared polygenic risk scores between cases and controls, burden between ND-CNV carriers and non-carriers, and employed two-sample Mendelian randomization to test whether genetic liability to cleft causally influences neurodevelopmental outcomes.

**Results:** Linkage disequilibrium score regression revealed little evidence of genetic correlations between CL/P and any of the eight traits examined. Within the cleft population, polygenic risk scores demonstrated expected associations with developmental and behavioural outcomes; however, children with CL/P did not have increased polygenic risk scores for ADHD, autism, depression, anxiety, schizophrenia, bipolar disorder, or lower scores for educational attainment or intelligence compared to controls. Mendelian randomization provided no robust evidence that genetic liability to cleft causally influences neurodevelopmental outcomes. ND-CNV carriers did not differ from non-carriers in polygenic burden.

**Conclusions:** The increased neurodevelopmental risk observed in CL/P does not appear to be explained by shared common genetic architecture with psychiatric disorders, contrasting with established rare variant contributions. Polygenic risk scores for neurodevelopmental traits predict behavioural outcomes within the cleft population similarly to the general population, indicating these genetic factors operate independently of cleft status but remain clinically relevant.

## Background

Cleft lip and/or palate (CL/P) is the most common craniofacial congenital anomaly, affecting approximately one in 1,000 live births globally [1]. Orofacial clefts encompass distinct phenotypes—cleft lip only (CLO), cleft palate only (CPO), and cleft lip with palate (CLP)— with evidence suggesting different aetiological pathways [2]. Genome-wide association studies have identified different genetic loci for cleft palate only compared to cleft lip with or without palate, supporting long-standing clinical observations of aetiological heterogeneity [3-6]. Approximately 30% of children with CL/P present with additional congenital anomalies or as part of a recognised genetic syndrome. This proportion is notably higher amongst those with CPO, where syndromic forms may account for over 50% of cases [7, 8].

Compared to children in the general population, children with CL/P are more likely to have neurodevelopmental difficulties, including autism spectrum disorder, Attention-Deficit Hyperactivity Disorder (ADHD) and educational challenges [9, 10]. Meta-analyses have also reported elevated risk for depression and anxiety, with effects persisting into adulthood, substantially impacting subsequent quality of life [10]. Additionally, having an orofacial cleft can also have an impact on educational attainment, with a lower academic performance among children with CLO or CLP compared to their peers [11]. While environmental factors secondary to CL/P—such as surgical interventions, speech/communication difficulties, and social experiences including bullying—might be a contributing factor [12], an emerging body of evidence suggests genetic factors may play a substantial role in the neurodevelopmental comorbidities [2]. However, observational studies can often fail to distinguish correlation from causation, as unmeasured confounding and reverse causation may bias observed associations. Mendelian randomization (MR) offers a complementary approach—using genetic variants as instrumental variables— to test whether genetic liability to cleft causally influences neurodevelopmental outcomes, circumventing many of the biases inherent to observational designs [13, 14].

Previous research has identified that neurodevelopmental copy number variants (ND-CNVs; rare genomic micro-deletions or duplications previously implicated in neurodevelopmental disorders) [15], are enriched in children with CL/P (3.7% vs 2.2% in controls), with CNV carriers showing delays in early development and higher rates of behavioural difficulties [2]. This enrichment was particularly pronounced in children born with CPO (5.1%); these children were three times more likely to carry ND-CNVs compared to those with CLO. Those with ND-CNVs were more likely to experience behavioural difficulties, including conduct, emotional, hyperactivity and peer problems, at levels indicating clinically significant impairment.

ND-CNVs contribute to neurodevelopmental comorbidities in cleft, but these rare genetic variants are only present in a small number of children with CL/P. This raises the question of whether common genetic variants shared with neurodevelopmental disorders might also contribute to these comorbidities, given the highly polygenic profile of many of these disorders/traits [16]. This can be addressed using polygenic risk scores [17], which are an important resource for investigating the contribution of common variants. Extensive genetic correlations exist across psychiatric and neurodevelopmental disorders, suggesting shared biological pathways [18]. Previous studies have found little evidence for interactions between ND-CNVs and polygenic risk scores in population cohorts [19, 20] though within clinical cohorts such as 22q11.2 deletion syndrome, polygenic scores for schizophrenia or cognitive ability can provide additional risk stratification [21].

The Cleft Collective provides a unique opportunity to investigate these questions through longitudinal assessments of neurodevelopmental outcomes. This UK-based national cohort study of children with CL/P includes detailed phenotypic characterisation at multiple timepoints from infancy through childhood, capturing the developmental trajectory of children born with cleft [22]. Combined with our previous findings of rare variant contributions [2], this enables a comprehensive examination of how both common and rare genetic variation contribute to neurodevelopmental risk in this population.

Using data from 2,313 genotyped children with CL/P from the Cleft Collective [22] and 7,913 population controls from the Millennium Cohort Study [23], we first tested whether CL/P shares common variant architecture with cognitive, neurodevelopmental and psychiatric traits using genome-wide genetic correlations. We subsequently investigated whether polygenic risk scores for neurodevelopmental/psychiatric traits and for cleft itself are associated with behavioural and developmental outcomes within children with CL/P across multiple timepoints, and examined whether children with CL/P carry elevated polygenic risk for these traits compared to controls, both overall and stratified by cleft subtype. To understand the relationship between common and rare genetic variation in individuals with CL/P, we compared cognitive, neurodevelopmental and psychiatric and cleft polygenic risk scores between ND-CNV carriers and non-carriers. Finally, we employed two-sample Mendelian randomization to test whether genetic liability to CL/P causally influences neurodevelopmental and psychiatric outcomes.

## Methods

### Study population

The Cleft Collective is an ongoing UK-based national cohort study of children born with CL/P, their parents, and siblings, with detailed recruitment and data collection procedures described elsewhere [22]. The current study included 2,313 children with genetic data passing quality controls. Controls comprised 7,913 children from the Millennium Cohort Study, a nationally representative UK birth cohort of children born in 2000-2001 [23]. The Cleft Collective received ethical approval from the South-West Central Bristol NRES Ethics Committee (REC 13/SW/0064). The data for the current project was requested through application CC048-ES.

### Genotyping and quality control

Biological samples from cases were genotyped using the Illumina Global Screening Array version 3 at Bristol Bioresource Laboratories. After initial quality control removing samples with call rates <95%, excess heterozygosity, or sex mismatches, we performed imputation using the Haplotype Reference Consortium panel. Post-imputation quality control excluded variants with INFO score <0.8, minor allele frequency <0.01, missing rate >0.1, or Hardy-Weinberg equilibrium midp <10^−6^ in control samples. The final dataset comprised 5,406,770 variants for analysis. Control genotype data underwent identical processing. Principal component analysis was performed to identify population outliers and derive covariates for association analyses. More details can be in found in the initial report of the Cleft Collective Genome-Wide association study [4].

### ND-CNV Calling

Biological samples underwent genotyping using the Illumina Global Screening Array version 3 at Bristol Bioresource Laboratories. We previously [2] implemented a comprehensive CNV detection pipeline focused on 54 pre-defined neurodevelopmental CNVs previously implicated in neurodevelopmental disorders. CNV detection was performed using PennCNV version 1.0.5, which employs a hidden Markov model incorporating log R ratio and B allele frequency data [24]. Initial quality control excluded samples with call rates below 95%, excess heterozygosity, or sex mismatches. All CNV calls underwent manual validation through visual inspection of probe-level data. Where parental samples were available, we determined inheritance status (inherited versus de novo) through comparative analysis. CNVs were classified as pathogenic based on established databases including DECIPHER [25] and ClinGen [26], with particular focus on recurrent neurodevelopmental CNVs including 15q11.2, 16p11.2, and 22q11.2 deletions and duplications.

### Genetic correlation analysis

We estimated genetic correlations between cleft and neurodevelopmental disorders using linkage disequilibrium score regression (LDSC) [27] implemented in the ldsc software package [28]. We used pre-computed LD scores from the 1000 Genomes European reference panel [29] and restricted analyses to HapMap3 [30] SNPs to ensure well-imputed variants. Multiple testing correction used false discovery rate with q-value threshold of 0.05.

### Polygenic risk score calculation

We calculated polygenic risk scores using PRSice-2 software[31] with a clumping and thresholding approach. Discovery GWAS summary statistics were obtained from the latest available studies: ADHD [32], autism spectrum disorder [33], schizophrenia [34], bipolar disorder [35], depression [36], anxiety disorders[37], educational attainment [38] and intelligence [39]. To derive cleft polygenic scores, we used an external meta-analysis of cleft GWAS that used primarily CLO and CLP samples that did not overlap with the Cleft Collective [11], to avoid overfitting. The score was calculated for 2,313 genotyped Cleft Collective cases and 7,913 population controls from the Millennium Cohort Study.

We applied standard clumping parameters across all generated polygenic risk scores for comparability (r^2^ = 0.1, 250kb window) and used a p-value threshold of 0.05 for primary analyses. Scores were standardized to mean 0 and standard deviation 1 in the combined sample.

### Mendelian randomization

We performed two-sample Mendelian randomization [13, 14] using genome-wide significant cleft variants from the Cleft Collective [4] as instrumental variables. MR relies on three core assumptions: (1) relevance (genetic variants associated with exposure), (2) independence (variants not associated with confounders), and (3) exclusion restriction (variants affect outcome only through exposure). We calculated F-statistics to assess instrument strength, where F = β^2^/SE^2^; mean F-statistic = 45.4 (range 33.1-98.8), with all values >10 indicating adequate instrument strength. Primary analyses used the inverse variance weighted (IVW) method. Sensitivity analyses included: MR-Egger regression with intercept test for directional pleiotropy; weighted median estimation (robust when up to 50% of instruments are invalid); and weighted mode estimation. We assessed heterogeneity using Cochran’s Q statistic and conducted leave-one-out analyses to identify the effect of influential variants. Analyses were implemented using the TwoSampleMR package in R[40]. Results were corrected for multiple testing using FDR and reported while consulting the STROBE-MR checklist[41]

### Statistical analysis

Case-control comparisons of polygenic risk scores were performed using logistic regression adjusting for sex and ten principal components. Within-case analyses of developmental outcomes used linear regression with the same covariates plus age at assessment where relevant. For cleft subtype analyses, we used multinomial logistic regression with cleft lip only as the reference category. To test independence of rare and common variant effects, we compared polygenic risk scores between ND-CNV carriers and non-carriers using logistic regression, with and without adjustment for cleft subtype.

### Measures

#### Cleft Type

Cleft phenotype classification followed International Classification of Diseases protocols with verification through clinical records and surgical notes. Primary categorisation distinguished between: A) cleft lip only (CLO), involving the lip with or without alveolar involvement but intact palate; B) cleft palate only CPO), involving hard and/or soft palate without lip involvement; and C) cleft lip and palate (CLP), involving both structures.

### Developmental, behavioural and mental health measures

#### Ages and Stages Questionnaire (ASQ-3)

The ASQ-3 [42] provided comprehensive developmental screening across five domains at 18 months, 3 years, and 5 years, completed by the parents of the Cleft Collective. The communication domain (6 items) assessed receptive and expressive language, including understanding of simple instructions, vocabulary size, and sentence complexity. The gross motor domain (6 items) evaluated large muscle movements and coordination, such as walking, running, jumping, and balance. Fine motor skills (6 items) measured hand-eye coordination, manipulation of small objects, and early writing skills. The problem-solving domain (6 items) assessed cognitive abilities including object permanence, cause-and-effect understanding, and early mathematical concepts. Personal-social development (6 items) evaluated self-help skills, social interaction, and emotional regulation. Each item was scored as “yes” (10 points), “sometimes” (5 points), or “not yet” (0 points), with domain scores ranging from 0-60. Lower scores indicate developmental concerns, with clinical cut-offs at 2 standard deviations below age-specific means.

#### Ages and Stages Questionnaire: Social-Emotional, Second Edition (ASQ:SE-2)

The ASQ:SE-2 [43] complements the ASQ-3 by providing detailed assessment of social-emotional competence and potential behavioural concerns at the same timepoints. This instrument contains age-specific item sets evaluating seven behavioural areas: self-regulation (ability to calm when upset, adapt to changes), compliance (following rules and instructions), adaptive functioning (daily living skills appropriate for age), autonomy (independence and self-initiation), affect (emotional expression and empathy), social communication (interaction with peers and adults), and interaction with people (relationship quality and attachment behaviours). Items are scored on a three-point scale with an additional “concern” marker for behaviours parents find concerning. Total scores vary by age form, with higher scores indicating greater social-emotional difficulties. Clinical cut-offs are empirically derived for each age, identifying children requiring further evaluation or intervention.

#### Strengths and Difficulties Questionnaire (SDQ)

The SDQ [44] provided standardized behavioural screening at ages 5, 8, and 10 years through 25 items across five subscales. The emotional symptoms scale (5 items) assessed internalising problems including anxiety, depression, and somatic complaints (e.g., “Often complains of headaches,” “Many worries”). The conduct problems scale (5 items) evaluated externalising behaviours including aggression and rule-breaking (e.g., “Often has temper tantrums,” “Generally obedient” - reverse scored). The hyperactivity/inattention scale (5 items) measured ADHD symptoms (e.g., “Restless, overactive,” “Easily distracted”). The peer relationship problems scale (5 items) assessed social difficulties (e.g., “Rather solitary,” “Picked on or bullied”). The prosocial behaviour scale (5 items) evaluated positive social behaviours (e.g., “Considerate of others,” “Shares readily”). Each item was rated as “not true” 0, “somewhat true” 1, or “certainly true” 2. A total difficulties score (0-40) was computed from the first four subscales. Subscales as well as the full SDQ measure were treated for the purposes of the analyses in this investigation as continuous scales. Both maternal and paternal ratings were collected where available; when both were present, we used the mean of the two to increase reliability.

#### Additional Mental Health Measures

At age 10, we administered supplementary validated instruments to capture specific mental health concerns which might emerge at this age group. The Mood and Feelings Questionnaire (MFQ) short version[45] contained 13 items assessing depressive symptoms over the past two weeks (e.g., “felt miserable or unhappy,” “felt lonely”), scored 0 (not true), 1 (sometimes), or 2 (true), with total scores ranging 0-26 and scores ≥12 suggesting clinical depression. The Screen for Child Anxiety Related Disorders (SCARED)[46, 47] included 41 items measuring anxiety symptoms across five factors: panic/somatic symptoms, generalized anxiety, separation anxiety, social anxiety, and school avoidance. Items were rated 0 (not true), 1 (somewhat true), or 2 (very true), with total scores ≥25 indicating potential anxiety disorder. These instruments provided dimensional assessment of mental health symptoms. Both scales were completed by the parents of the participants.

## Results

### Genome-wide genetic correlations using LD Score Regression

After FDR correction for multiple testing, we found little evidence of genetic correlations between CL/P and any of the eight cognitive, neurodevelopmental and psychiatric traits examined (Figure 1). All genetic correlation coefficients (rg) ranged from -0.05 to +0.15. The strongest uncorrected correlations were observed with autism spectrum disorder (rg = 0.15, SE = 0.12, p-uncorrected = 0.20) and schizophrenia (rg = 0.10, SE = 0.07, p-uncorrected = 0.17). [Figure 1 here]

**Figure 1:**
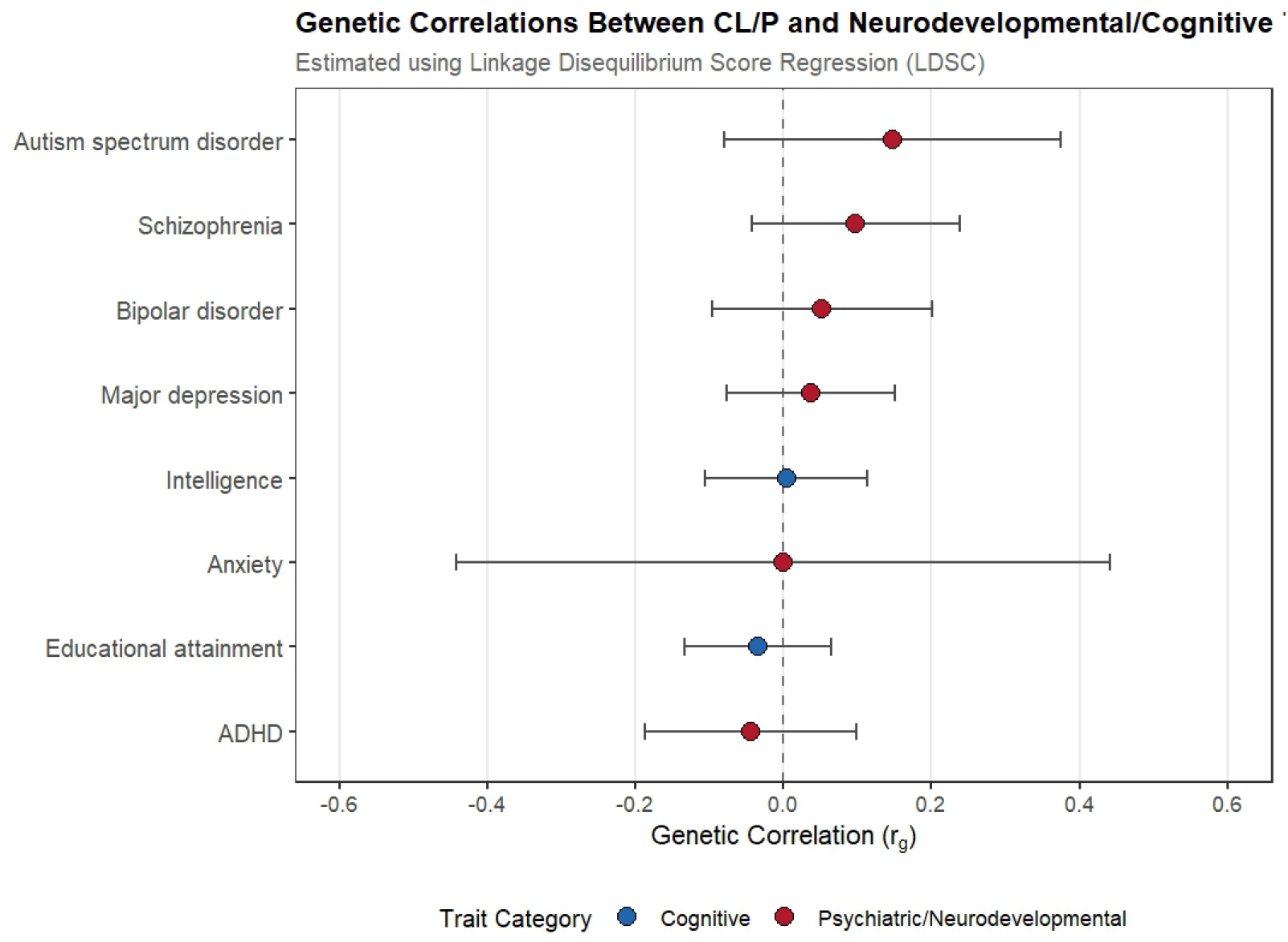
Genetic correlations between cleft lip/palate and cognitive, neurodevelopmental and psychiatric traits.

### Association of polygenic risk scores for cognitive, neurodevelopmental and psychiatric traits with developmental, behavioural and mental health difficulties measures within the Cleft Collective

We tested whether polygenic risk scores were associated with behavioural and developmental outcomes within children with CL/P. Sample sizes for each measure and timepoint are provided in Additional file Supplemental Table 1. Effect sizes (β) represent the change in outcome score per standard deviation increase in polygenic risk score.

### Ages and Stages Questionnaire (ASQ) Developmental Domains

Across ASQ developmental domains at 18 months, 3 years, and 5 years (higher scores = better performance), there was little evidence of consistent associations with any polygenic risk score (Table 1). Effect estimates were small, inconsistent in direction across timepoints, and confidence intervals were wide. No polygenic score demonstrated a consistent pattern of association with any ASQ developmental domain.

**Table 1.**
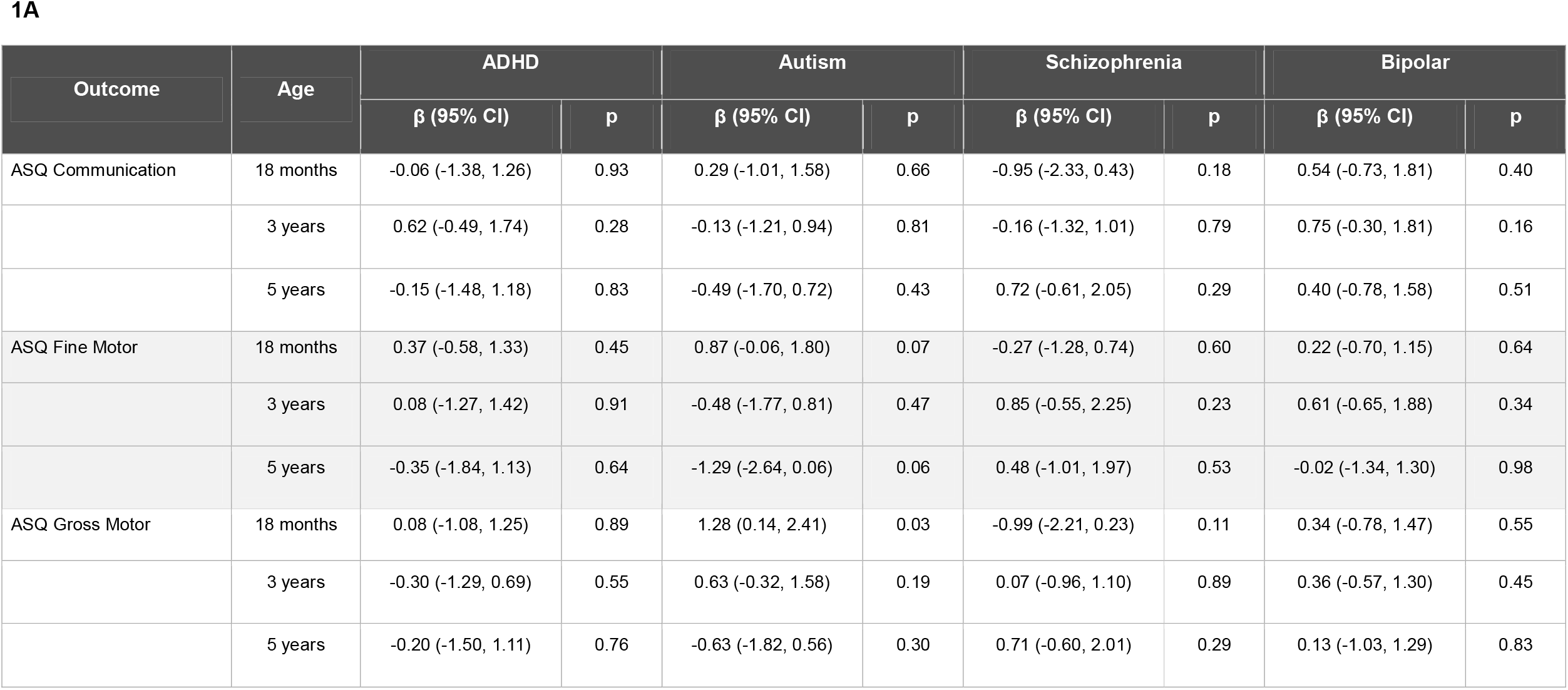
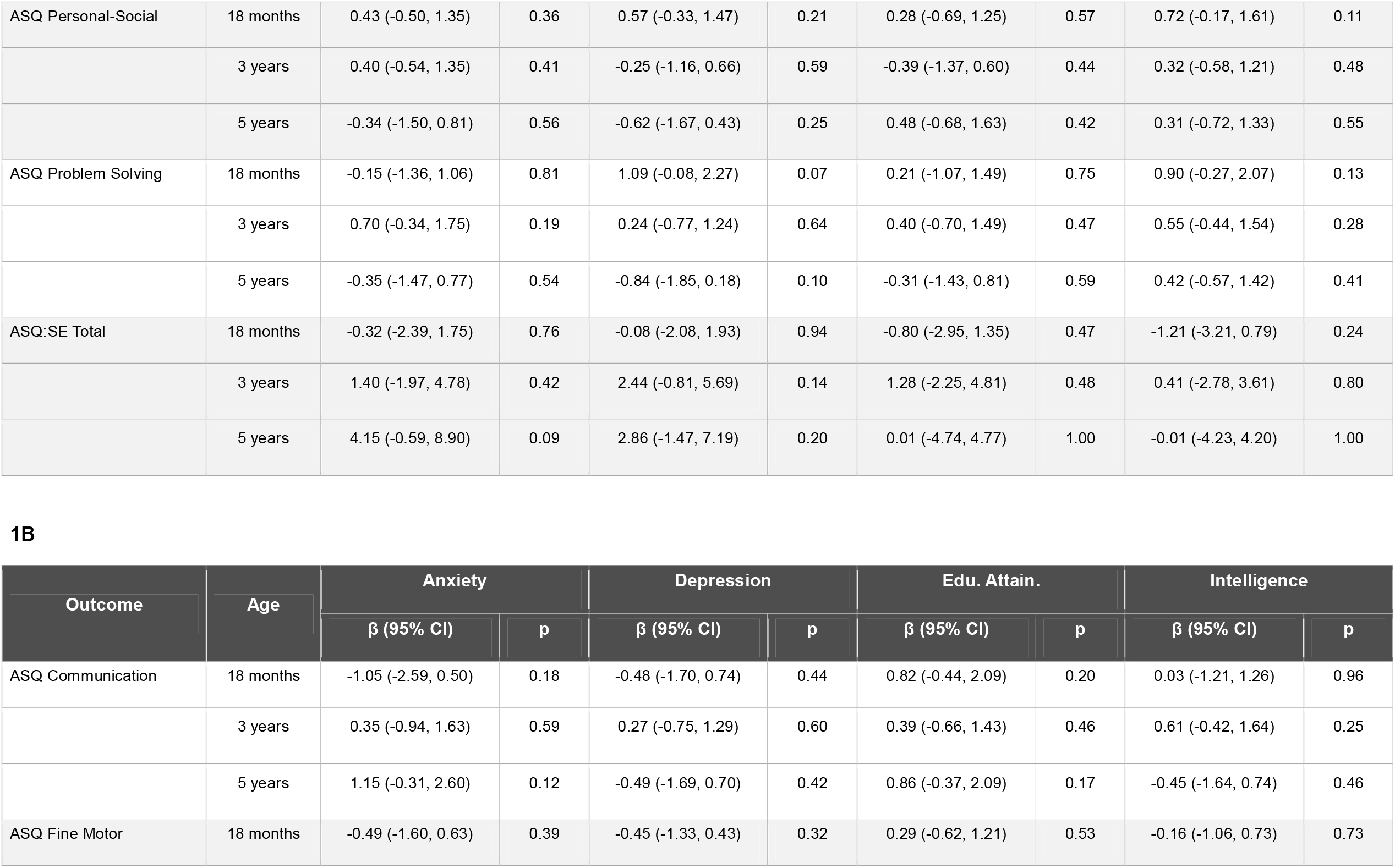
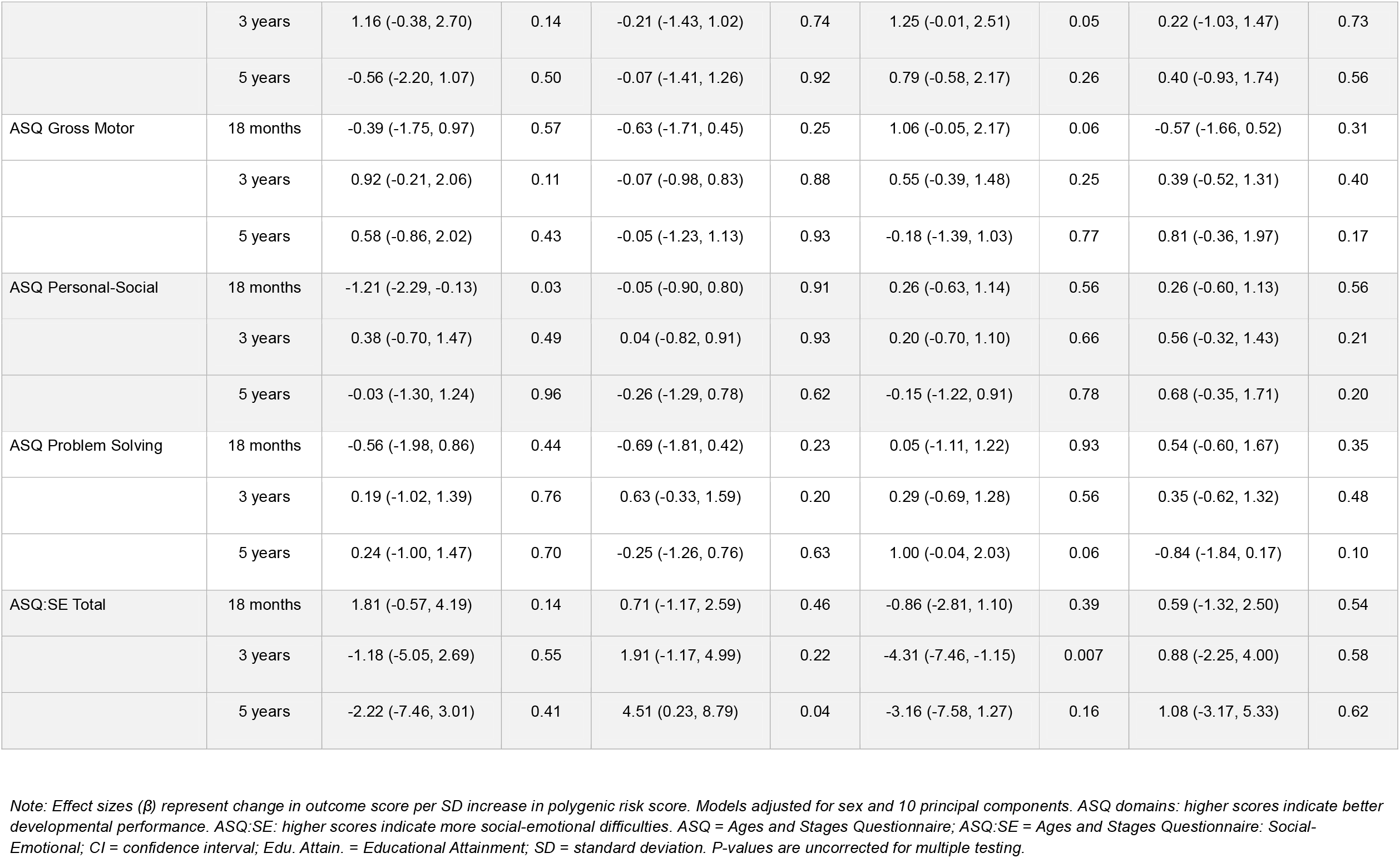
Association between polygenic risk scores for cognitive, neurodevelopmental and psychiatric traits and developmental outcomes (ASQ and ASQ:SE)

| Outcome | Age | ADHD |  | Autism |  | Schizophrenia |  | Bipolar |  |
| --- | --- | --- | --- | --- | --- | --- | --- | --- | --- |
| | | $\beta$ (95% CI) | p | $\beta$ (95% CI) | p | $\beta$ (95% CI) | p | $\beta$ (95% CI) | p |
| ASQ Communication | 18 months | -0.06 (-1.38, 1.26) | 0.93 | 0.29 (-1.01, 1.58) | 0.66 | -0.95 (-2.33, 0.43) | 0.18 | 0.54 (-0.73, 1.81) | 0.40 |
|  | 3 years | 0.62 (-0.49, 1.74) | 0.28 | -0.13 (-1.21, 0.94) | 0.81 | -0.16 (-1.32, 1.01) | 0.79 | 0.75 (-0.30, 1.81) | 0.16 |
|  | 5 years | -0.15 (-1.48, 1.18) | 0.83 | -0.49 (-1.70, 0.72) | 0.43 | 0.72 (-0.61, 2.05) | 0.29 | 0.40 (-0.78, 1.58) | 0.51 |
| ASQ Fine Motor | 18 months | 0.37 (-0.58, 1.33) | 0.45 | 0.87 (-0.06, 1.80) | 0.07 | -0.27 (-1.28, 0.74) | 0.60 | 0.22 (-0.70, 1.15) | 0.64 |
|  | 3 years | 0.08 (-1.27, 1.42) | 0.91 | -0.48 (-1.77, 0.81) | 0.47 | 0.85 (-0.55, 2.25) | 0.23 | 0.61 (-0.65, 1.88) | 0.34 |
|  | 5 years | -0.35 (-1.84, 1.13) | 0.64 | -1.29 (-2.64, 0.06) | 0.06 | 0.48 (-1.01, 1.97) | 0.53 | -0.02 (-1.34, 1.30) | 0.98 |
| ASQ Gross Motor | 18 months | 0.08 (-1.08, 1.25) | 0.89 | 1.28 (0.14, 2.41) | 0.03 | -0.99 (-2.21, 0.23) | 0.11 | 0.34 (-0.78, 1.47) | 0.55 |
|  | 3 years | -0.30 (-1.29, 0.69) | 0.55 | 0.63 (-0.32, 1.58) | 0.19 | 0.07 (-0.96, 1.10) | 0.89 | 0.36 (-0.57, 1.30) | 0.45 |
|  | 5 years | -0.20 (-1.50, 1.11) | 0.76 | -0.63 (-1.82, 0.56) | 0.30 | 0.71 (-0.60, 2.01) | 0.29 | 0.13 (-1.03, 1.29) | 0.83 |
| ASQ Personal-Social | 18 months | 0.43 (-0.50, 1.35) | 0.36 | 0.57 (-0.33, 1.47) | 0.21 | 0.28 (-0.69, 1.25) | 0.57 | 0.72 (-0.17, 1.61) | 0.11 |
|  | 3 years | 0.40 (-0.54, 1.35) | 0.41 | -0.25 (-1.16, 0.66) | 0.59 | -0.39 (-1.37, 0.60) | 0.44 | 0.32 (-0.58, 1.21) | 0.48 |
|  | 5 years | -0.34 (-1.50, 0.81) | 0.56 | -0.62 (-1.67, 0.43) | 0.25 | 0.48 (-0.68, 1.63) | 0.42 | 0.31 (-0.72, 1.33) | 0.55 |
| ASQ Problem Solving | 18 months | -0.15 (-1.36, 1.06) | 0.81 | 1.09 (-0.08, 2.27) | 0.07 | 0.21 (-1.07, 1.49) | 0.75 | 0.90 (-0.27, 2.07) | 0.13 |
|  | 3 years | 0.70 (-0.34, 1.75) | 0.19 | 0.24 (-0.77, 1.24) | 0.64 | 0.40 (-0.70, 1.49) | 0.47 | 0.55 (-0.44, 1.54) | 0.28 |
|  | 5 years | -0.35 (-1.47, 0.77) | 0.54 | -0.84 (-1.85, 0.18) | 0.10 | -0.31 (-1.43, 0.81) | 0.59 | 0.42 (-0.57, 1.42) | 0.41 |
| ASQ:SE Total | 18 months | -0.32 (-2.39, 1.75) | 0.76 | -0.08 (-2.08, 1.93) | 0.94 | -0.80 (-2.95, 1.35) | 0.47 | -1.21 (-3.21, 0.79) | 0.24 |
|  | 3 years | 1.40 (-1.97, 4.78) | 0.42 | 2.44 (-0.81, 5.69) | 0.14 | 1.28 (-2.25, 4.81) | 0.48 | 0.41 (-2.78, 3.61) | 0.80 |
|  | 5 years | 4.15 (-0.59, 8.90) | 0.09 | 2.86 (-1.47, 7.19) | 0.20 | 0.01 (-4.74, 4.77) | 1.00 | -0.01 (-4.23, 4.20) | 1.00 |

1B
| Outcome | Age | Anxiety |  | Depression |  | Edu. Attain. |  | Intelligence |  |
| --- | --- | --- | --- | --- | --- | --- | --- | --- | --- |
| | | $\beta$ (95% CI) | p | $\beta$ (95% CI) | p | $\beta$ (95% CI) | p | $\beta$ (95% CI) | p |
| ASQ Communication | 18 months | -1.05 (-2.59, 0.50) | 0.18 | -0.48 (-1.70, 0.74) | 0.44 | 0.82 (-0.44, 2.09) | 0.20 | 0.03 (-1.21, 1.26) | 0.96 |
|  | 3 years | 0.35 (-0.94, 1.63) | 0.59 | 0.27 (-0.75, 1.29) | 0.60 | 0.39 (-0.66, 1.43) | 0.46 | 0.61 (-0.42, 1.64) | 0.25 |
|  | 5 years | 1.15 (-0.31, 2.60) | 0.12 | -0.49 (-1.69, 0.70) | 0.42 | 0.86 (-0.37, 2.09) | 0.17 | -0.45 (-1.64, 0.74) | 0.46 |
| ASQ Fine Motor | 18 months | -0.49 (-1.60, 0.63) | 0.39 | -0.45 (-1.33, 0.43) | 0.32 | 0.29 (-0.62, 1.21) | 0.53 | -0.16 (-1.06, 0.73) | 0.73 |
|  | 3 years | 1.16 (-0.38, 2.70) | 0.14 | -0.21 (-1.43, 1.02) | 0.74 | 1.25 (-0.01, 2.51) | 0.05 | 0.22 (-1.03, 1.47) | 0.73 |
|  | 5 years | -0.56 (-2.20, 1.07) | 0.50 | -0.07 (-1.41, 1.26) | 0.92 | 0.79 (-0.58, 2.17) | 0.26 | 0.40 (-0.93, 1.74) | 0.56 |
| ASQ Gross Motor | 18 months | -0.39 (-1.75, 0.97) | 0.57 | -0.63 (-1.71, 0.45) | 0.25 | 1.06 (-0.05, 2.17) | 0.06 | -0.57 (-1.66, 0.52) | 0.31 |
|  | 3 years | 0.92 (-0.21, 2.06) | 0.11 | -0.07 (-0.98, 0.83) | 0.88 | 0.55 (-0.39, 1.48) | 0.25 | 0.39 (-0.52, 1.31) | 0.40 |
|  | 5 years | 0.58 (-0.86, 2.02) | 0.43 | -0.05 (-1.23, 1.13) | 0.93 | -0.18 (-1.39, 1.03) | 0.77 | 0.81 (-0.36, 1.97) | 0.17 |
| ASQ Personal-Social | 18 months | -1.21 (-2.29, -0.13) | 0.03 | -0.05 (-0.90, 0.80) | 0.91 | 0.26 (-0.63, 1.14) | 0.56 | 0.26 (-0.60, 1.13) | 0.56 |
|  | 3 years | 0.38 (-0.70, 1.47) | 0.49 | 0.04 (-0.82, 0.91) | 0.93 | 0.20 (-0.70, 1.10) | 0.66 | 0.56 (-0.32, 1.43) | 0.21 |
|  | 5 years | -0.03 (-1.30, 1.24) | 0.96 | -0.26 (-1.29, 0.78) | 0.62 | -0.15 (-1.22, 0.91) | 0.78 | 0.68 (-0.35, 1.71) | 0.20 |
| ASQ Problem Solving | 18 months | -0.56 (-1.98, 0.86) | 0.44 | -0.69 (-1.81, 0.42) | 0.23 | 0.05 (-1.11, 1.22) | 0.93 | 0.54 (-0.60, 1.67) | 0.35 |
|  | 3 years | 0.19 (-1.02, 1.39) | 0.76 | 0.63 (-0.33, 1.59) | 0.20 | 0.29 (-0.69, 1.28) | 0.56 | 0.35 (-0.62, 1.32) | 0.48 |
|  | 5 years | 0.24 (-1.00, 1.47) | 0.70 | -0.25 (-1.26, 0.76) | 0.63 | 1.00 (-0.04, 2.03) | 0.06 | -0.84 (-1.84, 0.17) | 0.10 |
| ASQ:SE Total | 18 months | 1.81 (-0.57, 4.19) | 0.14 | 0.71 (-1.17, 2.59) | 0.46 | -0.86 (-2.81, 1.10) | 0.39 | 0.59 (-1.32, 2.50) | 0.54 |
|  | 3 years | -1.18 (-5.05, 2.69) | 0.55 | 1.91 (-1.17, 4.99) | 0.22 | -4.31 (-7.46, -1.15) | 0.007 | 0.88 (-2.25, 4.00) | 0.58 |
|  | 5 years | -2.22 (-7.46, 3.01) | 0.41 | 4.51 (0.23, 8.79) | 0.04 | -3.16 (-7.58, 1.27) | 0.16 | 1.08 (-3.17, 5.33) | 0.62 |

*Note: Effect sizes ( $\beta$ ) represent change in outcome score per SD increase in polygenic risk score. Models adjusted for sex and 10 principal components. ASQ domains: higher scores indicate better developmental performance. ASQ:SE: higher scores indicate more social-emotional difficulties. ASQ = Ages and Stages Questionnaire; ASQ:SE = Ages and Stages Questionnaire: Social-Emotional; CI = confidence interval; Edu. Attain. = Educational Attainment; SD = standard deviation. P-values are uncorrected for multiple testing.*

### Ages and Stages Questionnaire: Social-Emotional (ASQ:SE)

For social-emotional difficulties on the ASQ:SE (higher scores indicate more difficulties), higher educational attainment polygenic score was consistently associated with fewer social-emotional difficulties across all timepoints [18 months (β=-0.86, 95% CI: -2.81, 1.10), 3 years (β=-4.31, 95% CI: -7.46, -1.15), and 5 years (β=-3.16, 95% CI: -7.58, 1.27)]. Depression polygenic score showed a pattern of increasing association with social-emotional difficulties across childhood, with effect estimates increasing with age (18 months (β=0.71, 95% CI: - 1.17, 2.59), 3 years (β=1.91, 95% CI: -1.17, 4.99), and 5 years (β=4.51, 95% CI: 0.23, 8.79)).

### Strengths and Difficulties Questionnaire (SDQ)

Associations between polygenic scores and SDQ outcomes are presented in Table 2. There was evidence of the ADHD polygenic risk score being associated with higher SDQ total difficulties score at all timepoints and effect estimates increased in magnitude with age: 5 years (β=0.81, 95% CI: 0.20, 1.43), 8 years (β=1.37, 95% CI: 0.35, 2.40), and 10 years (β=1.62, 95% CI: 0.53, 2.72). This pattern was mirrored across SDQ subscales (similar estimate effects and direction for the conduct problems, hyperactivity and peer problems scales). Educational attainment polygenic risk score demonstrated a pattern of protective associations with SDQ outcomes that complemented the ASQ:SE findings. Effect estimates for total difficulties were negative at all timepoints, with the association most precisely estimated at age 10 (β=-1.12, 95% CI: -2.09 to -0.15), with similar protective patterns emerging in the hyperactivity and conduct subscales. Finally, autism polygenic risk score also showed a consistent pattern of association with reduced prosocial behaviour at all timepoints (5 years: β=-0.20, 95% CI: -0.39, 0.00; 8 years: β=-0.24, 95% CI: -0.52, 0.05; 10 years: β=-0.27, 95% CI: -0.60, 0.05).

**Table 2.**
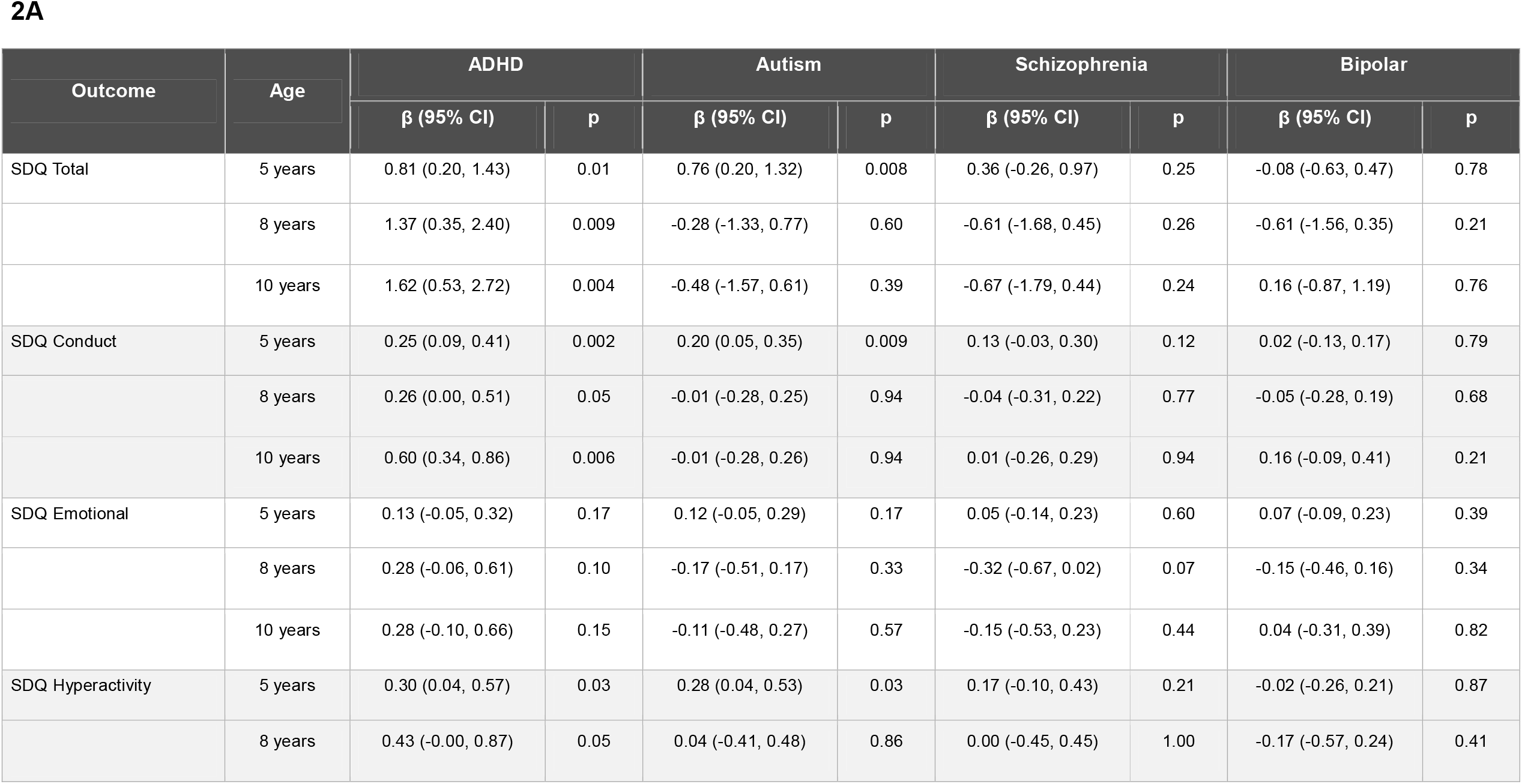
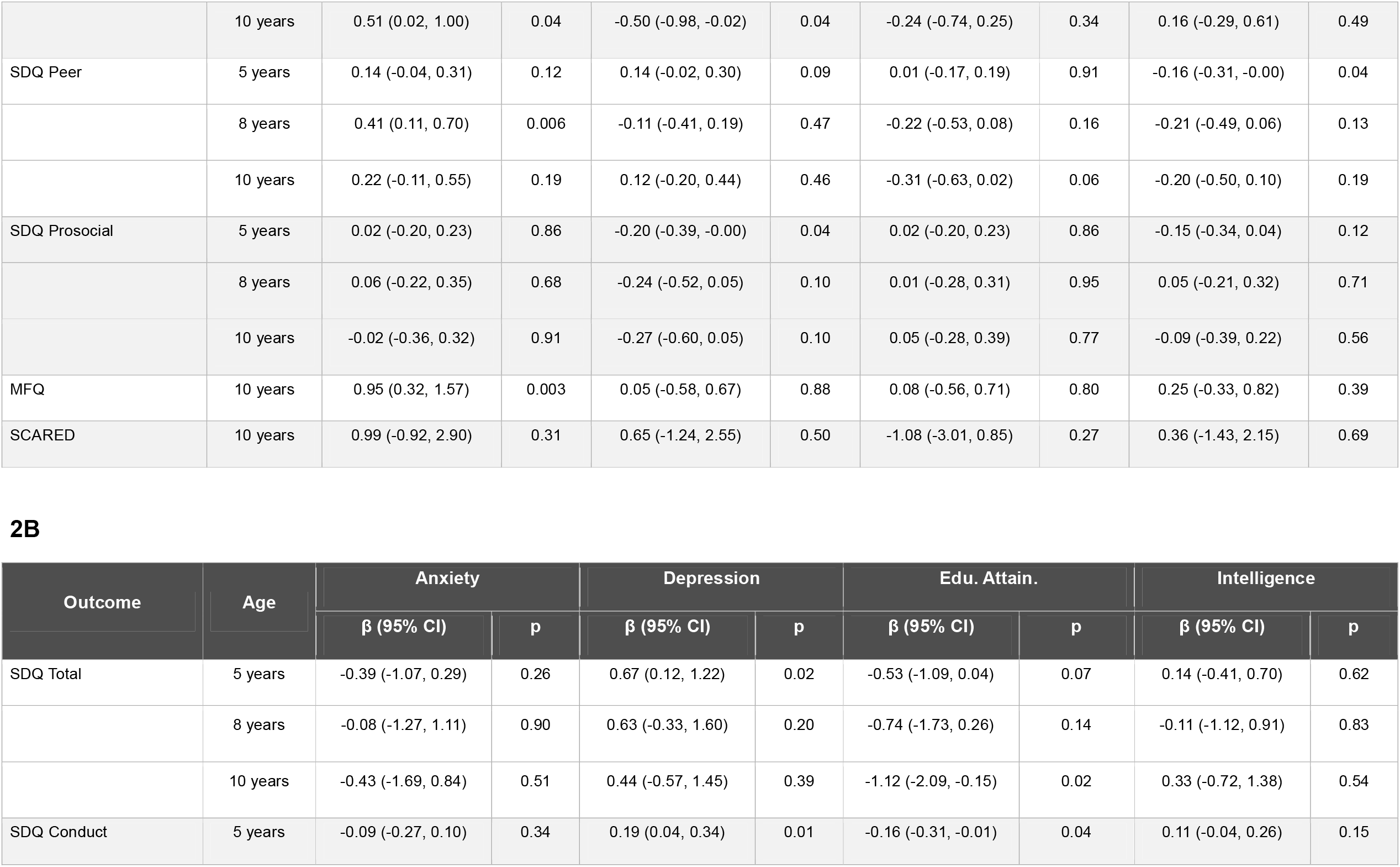
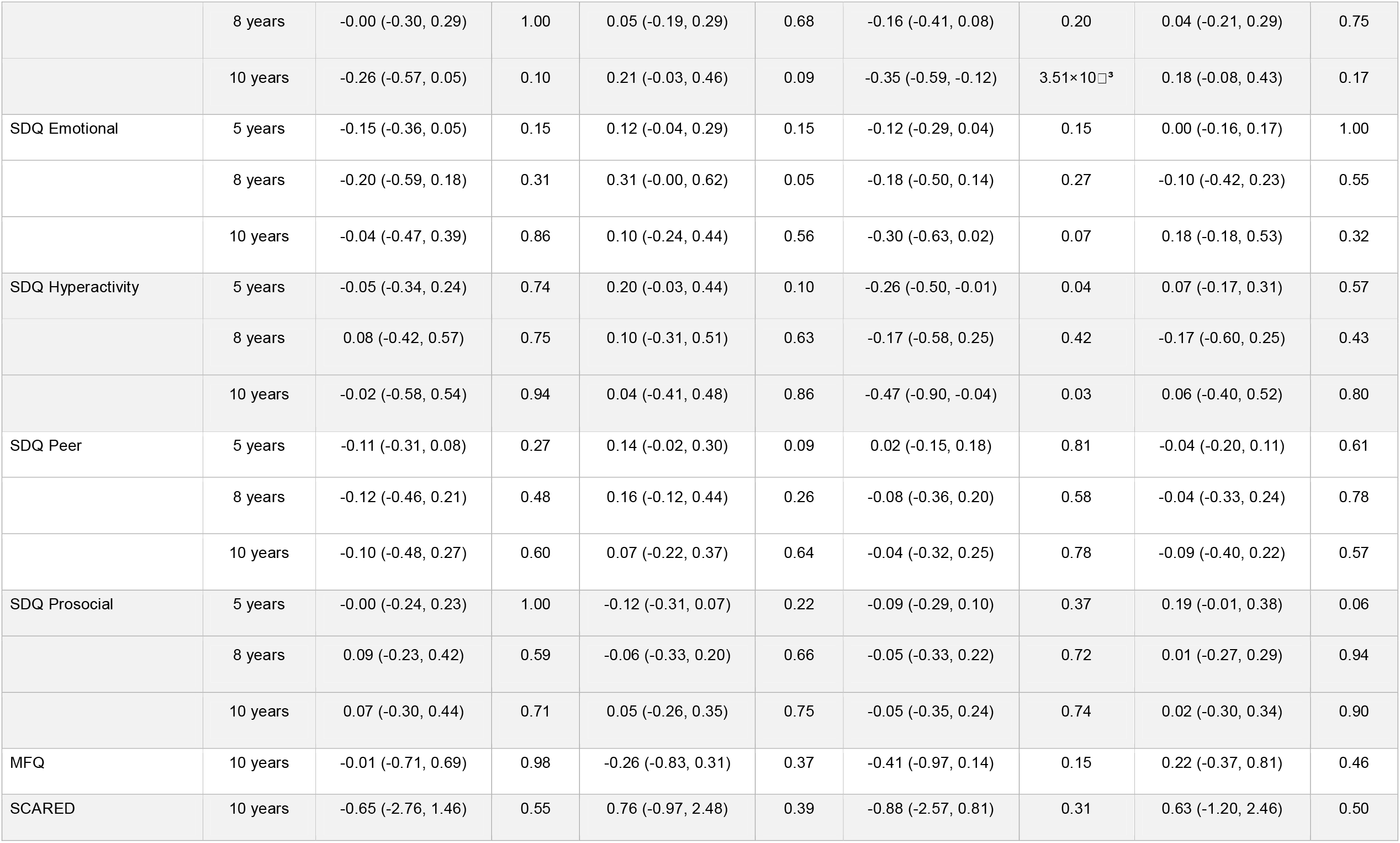
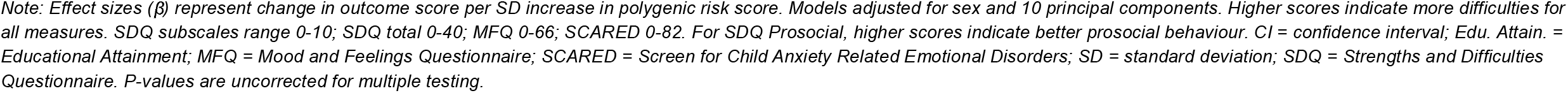
Association between polygenic risk scores for cognitive, neurodevelopmental and psychiatric traits and behavioural and mental health outcomes (SDQ, MFQ, SCARED)

### Mood and Feelings Questionnaire (MFQ)

At age 10 years, ADHD polygenic risk score was associated with higher depressive symptoms (β=0.95, 95% CI: 0.32, 1.57, p-uncorrected = 3×10^-3^). Depression polygenic risk score showed no evidence of association with MFQ (β=-0.26, 95% CI: -0.83, 0.31, p-uncorrected = 0.36) (Table 2).

### Screen for Child Anxiety Related Emotional Disorders (SCARED)

There was little evidence of association between any polygenic score and anxiety symptoms at age 10. Confidence intervals were wide for all exposures, limiting precision. Notably, anxiety polygenic risk score showed no association with SCARED scores (β=-0.65, 95% CI: - 2.76 to 1.46), with the point estimate in the opposite direction to expectation (Table 2).

### Non-syndromic Cleft Lip with or without palate polygenic score validation

The non-syndromic cleft lip with or without palate polygenic risk score (calculated from an external cleft cohort to avoid overfitting[11]) effectively discriminated cases from controls (OR = 1.35, 95% CI: 1.29, 1.42, p = 7×10^−37^), with particularly strong performance for CLO versus controls (OR = 1.70, p = 1.4×10^−31^) and CLP versus controls (OR = 1.60, p = 3.1×10^−39^), but as expected did not effectively differentiate CPO from controls (OR = 0.93, p = 0.06).

Additionally, we tested associations of the non-syndromic cleft lip with or without palate polygenic risk score with behavioural outcomes measured using the SDQ (ages 5, 8, 10 years), ASQ and ASQ-SE (18 months, 3 years, 5 years), MFQ (age 10), and SCARED (age 10) within children with CL/P. This polygenic risk score showed little evidence of consistent associations with behavioural and mental health outcomes across timepoints (Additional File, Supplemental Table 2).

### Polygenic risk scores for cognitive neurodevelopmental and psychiatric traits in cleft cases versus controls

Despite the observed associations between polygenic risk scores and behavioural outcomes within the cleft population, we found little evidence that children with CL/P carried differential polygenic risk compared to controls for any cognitive, neurodevelopmental or psychiatric trait examined (Table 3), with all odds ratios ranging from 0.91 to 1.03.

**Table 3:** Polygenic risk scores for cognitive, neurodevelopmental and psychiatric traits in children with cleft lip/palate compared to population controls.

| Polygenic Risk Score | OR (95% CI) | p-value |
| --- | --- | --- |
| ADHD | 0.97 (0.92, 1.03) | 0.32 |
| Depression | 1.02 (0.97, 1.08) | 0.43 |
| Anxiety | 1 (0.94, 1.06) | 0.99 |
| Schizophrenia | 0.96 (0.9, 1.02) | 0.17 |
| Autism | 1.01 (0.97, 1.07) | 0.58 |
| Bipolar disorder | 0.98 (0.93, 1.03) | 0.38 |
| Intelligence | 1.03 (0.99, 1.09) | 0.18 |
| Educational attainment | 0.96 (0.9, 1) | 0.07 |

Comparison of standardized polygenic risk scores between 2,313 children with cleft lip/palate from the Cleft Collective and 7,913 controls from the Millennium Cohort Study. Odds ratios (OR) represent the change in odds of having cleft per standard deviation increase in polygenic risk score, adjusted for biological sex and ten principal components. CI, confidence interval. P-values are uncorrected for multiple testing. Polygenic risk scores were calculated using $p < 0.05$ threshold from discovery GWAS.

### Association of polygenic risk scores for cognitive, neurodevelopmental and psychiatric traits with cleft subtypes

When stratifying by cleft subtype (CLO, CPO and CLP), we found minimal evidence for differential polygenic risk scores for neurodevelopmental disorders and cognitive traits across subtypes (Additional File, Supplemental Table 3). Anxiety polygenic risk score showed nominally lower scores in CLO compared to controls (OR=0.89, 95% CI: 0.79-1.00, p-uncorrected=0.046) and compared to CPO (OR=0.82, 95% CI: 0.71-0.95, p-uncorrected=0.009), while depression polygenic risk score was nominally higher in children with CLP compared to controls (OR=1.09, 95% CI: 1.01-1.18, p-uncorrected=0.026). Neither finding survived correction for multiple testing.

### Association of polygenic risk scores for cognitive, neurodevelopmental and psychiatric traits and Cleft with rare ND-CNVs

We also compared polygenic risk scores for cognitive, neurodevelopmental and psychiatric traits and cleft between children born with both CL/P and an ND-CNV (n=77) and children born with CL/P but not an ND-CNV (n=2,103) within the Cleft Collective. We found very little evidence to suggest differences in polygenic risk scores between the two groups for most traits (Table 4). The strongest signal was for educational attainment, where ND-CNV carriers showed a trend toward lower polygenic scores compared to non-carriers (OR=0.78, 95% CI: 0.62-1.00, p=0.05).

**Table 4.**
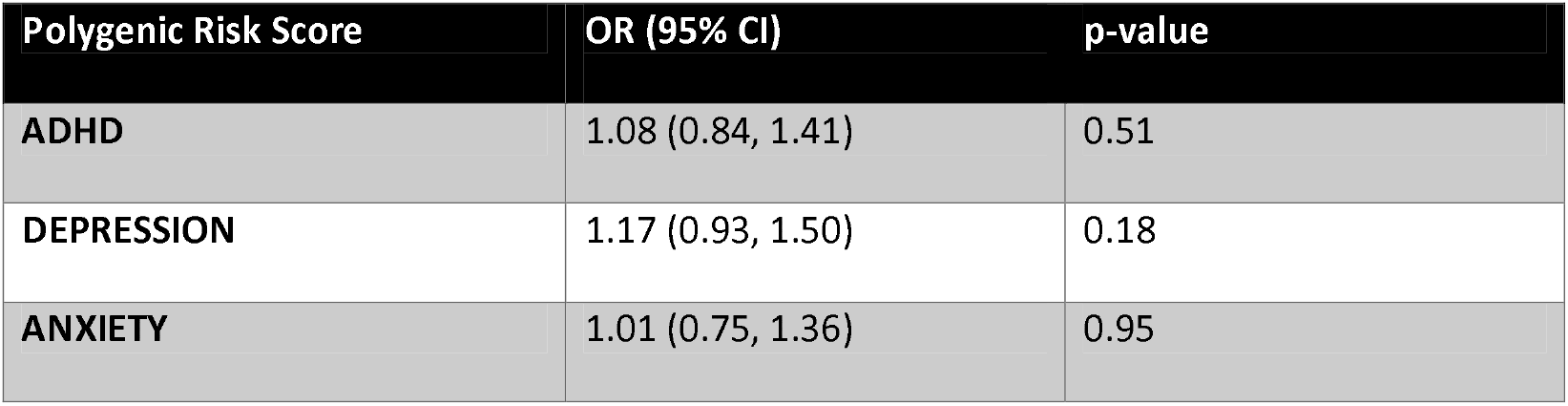
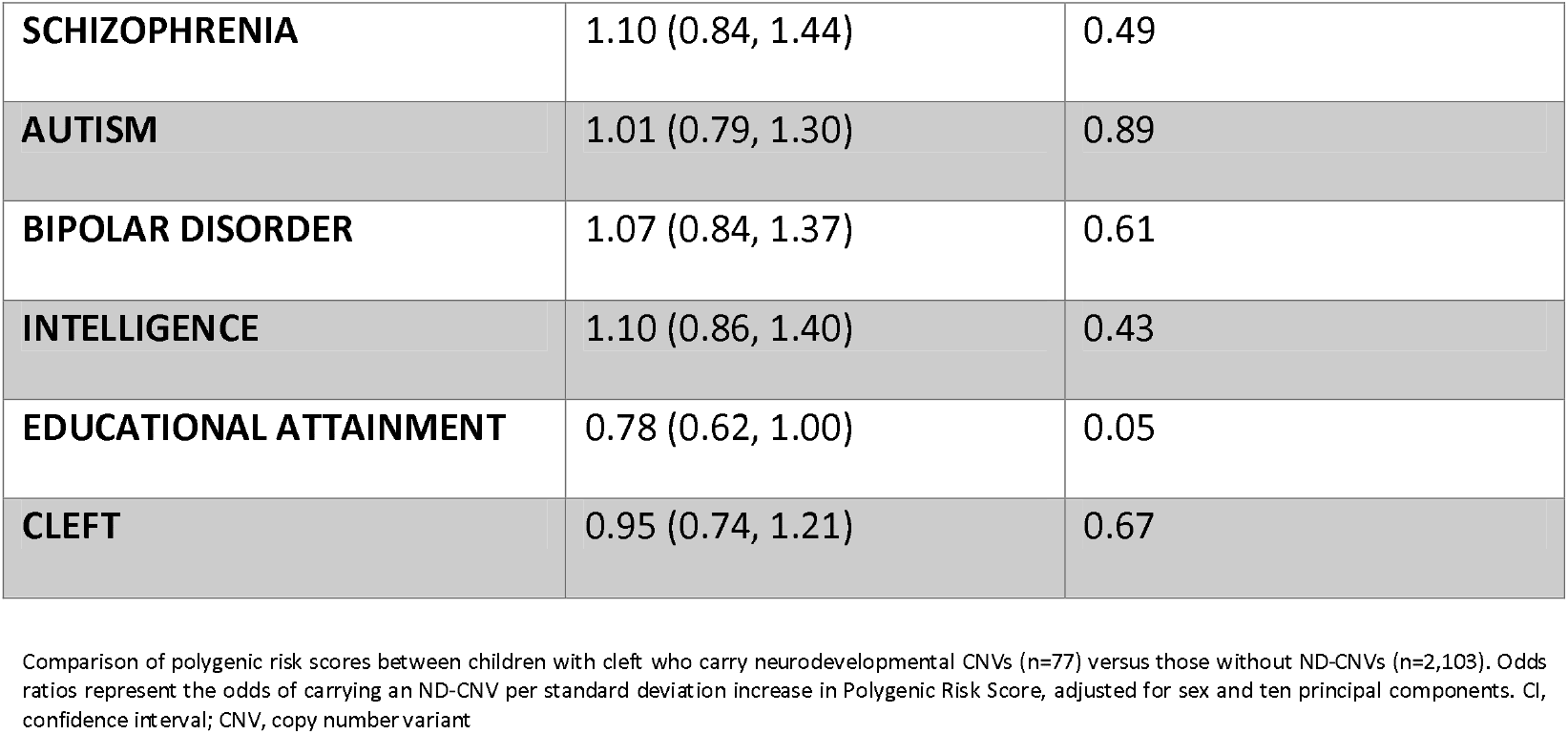
Polygenic Scores for cognitive, neurodevelopmental and psychiatric traits and cleft in ND-CNV carriers versus non-carriers among children with cleft.

| Polygenic Risk Score | OR (95% CI) | p-value |
| --- | --- | --- |
| ADHD | 1.08 (0.84, 1.41) | 0.51 |
| DEPRESSION | 1.17 (0.93, 1.50) | 0.18 |
| ANXIETY | 1.01 (0.75, 1.36) | 0.95 |
| **SCHIZOPHRENIA** | 1.10 (0.84, 1.44) | 0.49 |
| **AUTISM** | 1.01 (0.79, 1.30) | 0.89 |
| **BIPOLAR DISORDER** | 1.07 (0.84, 1.37) | 0.61 |
| **INTELLIGENCE** | 1.10 (0.86, 1.40) | 0.43 |
| **EDUCATIONAL ATTAINMENT** | 0.78 (0.62, 1.00) | 0.05 |
| **CLEFT** | 0.95 (0.74, 1.21) | 0.67 |

Comparison of polygenic risk scores between children with cleft who carry neurodevelopmental CNVs (n=77) versus those without ND-CNVs (n=2,103). Odds ratios represent the odds of carrying an ND-CNV per standard deviation increase in Polygenic Risk Score, adjusted for sex and ten principal components. CI, confidence interval; CNV, copy number variant

### Mendelian randomization of cleft genetic liability on cognitive, neurodevelopmental and psychiatric outcomes

We employed two-sample Mendelian randomization to test whether genetic liability to cleft causally influences differential risk of cognitive, neurodevelopmental and psychiatric traits. Using eight genome-wide significant cleft variants from the Cleft Collective all-cleft GWAS [4] as instrumental variables (mean F-statistic = 45.4, range 33.1-98.8, all >10 indicating adequate instrument strength, Additional File - Supplemental Table 4), we tested for causal effects on eight outcomes using summary statistics from the largest GWAS available (ADHD [32], autism [33], schizophrenia [34], bipolar disorder [48], depression [36], anxiety [37], educational attainment [38], and intelligence [39].

After FDR correction for multiple testing, we found very little evidence for causal effects of cleft genetic liability on any of the outcomes investigated (Additional File, Supplemental Table 5 and Supplemental Figure 1). The strongest evidence of causal effect was for autism (IVW β = 0.048, SE = 0.021, p = 0.026), though this did not survive FDR correction and showed no support from sensitivity analyses (weighted median p = 0.207, weighted mode p= 0.397).

MR-Egger intercept tests revealed evidence of directional pleiotropy only for depression (intercept = -0.018, p = 0.019). Accordingly, we report the MR-Egger slope estimate for depression (β = 0.070, SE = 0.021, p = 0.017), which differs substantially from the null IVW estimate (β = 0.004, p = 0.610), suggesting potential bias in the primary analysis for this outcome. For all other outcomes, MR-Egger intercepts (all p > 0.12), did not indicate evidence of directional pleiotropy.

Cochran’s Q tests revealed evidence of heterogeneity for bipolar disorder (Q = 22.0, p = 0.003), educational attainment (Q = 20.9, p = 0.002), depression (Q = 16.1, p = 0.024), and intelligence (Q = 22.2, p = 0.001). Leave-one-out analyses identified rs10172734 as driving heterogeneity for educational attainment and intelligence, and rs28361060 as driving heterogeneity for bipolar disorder (Additional File, Supplemental Table 6). Removal of these influential variants did not substantially change the findings.

## Discussion

In this comprehensive investigation of common genetic variant contributions to cognitive, neurodevelopmental and psychiatric traits in children with CL/P, we did not identify evidence of shared common variant genetic architecture between cleft and those traits. Using data from 2,313 children with CL/P from the Cleft Collective compared to 7,913 population controls from the Millennium Cohort Study, we did not observe genetic correlations between cleft and eight neurodevelopmental and cognitive traits examined through linkage disequilibrium score regression. Despite this, polygenic risk scores for these traits demonstrated expected associations with behavioural outcomes within the cleft population, indicating the scores function as valid instruments in this sample. Nevertheless, children with CL/P did not carry increased polygenic scores for ADHD, autism spectrum disorder, depression, anxiety, schizophrenia, bipolar disorder, or reduced polygenic load for educational attainment or intelligence compared to controls. This lack of differential polygenic burden extended across cleft subtypes, with no differences observed between children with CPO, CLO or CLP, despite the known aetiological heterogeneity and higher neurodevelopmental comorbidity in CPO. In contrast to the within-case findings, higher cleft polygenic risk score was not associated with increased behavioural difficulties within the Cleft Collective. Carriers of rare ND-CNVs did not have a differential common variant burden (either for cleft or for neurodevelopmental disorders and cognitive outcomes) compared to those without ND-CNV. Finally, Mendelian randomization analyses provided no robust evidence for causal effects of genetic liability to cleft on neurodevelopmental disorders and cognitive outcomes.

The absence of evidence for shared common variant architecture between cleft and cognitive, neurodevelopmental and psychiatric traits stands in contrast to our previous findings of enriched rare neurodevelopmental CNVs in children with CL/P [2]. This apparent dissociation may reflect the difference in biological pathways involved in cognitive, neurodevelopmental and psychiatric traits compared to cleft. Genome-wide association studies of cognitive, neurodevelopmental traits and psychiatric disorders consistently implicate neuronal biology, particularly synaptic structure and function, neurotransmission, and neurodevelopmental processes [49, 50]. Comprehensive enrichment analyses of neurological and psychiatric GWAS have demonstrated that psychiatric disorders specifically and consistently implicate neuronal signalling pathways [51] , as do ND-CNV studies [52, 53]. In contrast, GWAS of non-syndromic cleft lip with or without palate have identified genes involved in craniofacial development pathways, neural crest development and facial morphology [4, 54] but are distinct from the synaptic and neuronal pathways implicated in psychiatric disorders. The association observed between cleft and ND-CNVs may therefore reflect the larger phenotypic consequences these genomic rearrangements, which typically disrupt multiple genes and regulatory elements. This is also consistent with the observation that ND-CNV enrichment is most pronounced in cleft palate only, where syndromic forms— often involving broader developmental disruption—account for over 50% of presentations [7].

While statistical power was a consideration — the cleft polygenic score used in this study was derived from an earlier meta-analysis [11] to avoid sample overlap with the Cleft Collective and captures only a proportion of variance in cleft liability - — it is worth noting that this same cleft polygenic risk score effectively discriminated cases from controls, suggesting the methodology had adequate sensitivity to detect shared genetic signal where it exists. Additionally, across both LDSC and polygenic risk score analyses, the relatively small upper bounds of confidence intervals and odds ratios indicate that any shared risk, if present, would be quite small.

Polygenic risk scores for cognitive, neurodevelopmental traits and psychiatric conditions did demonstrate meaningful associations with behavioural outcomes within children with CL/P, consistent with patterns previously reported in the general population. ADHD polygenic risk score showed the most robust and consistent associations, predicting total difficulties scores across all three SDQ timepoints with effect sizes that increased with age, as well as conduct problems, hyperactivity, and depressive symptoms at age 10. These findings align with evidence from population-based cohorts demonstrating that ADHD polygenic risk is associated with ADHD symptoms, negative cognitive and educational outcomes, and conduct symptomatology [55]. Longitudinal studies have similarly shown that higher ADHD polygenic risk score predicts elevated hyperactivity/impulsivity and inattention from early childhood through adolescence [56] and has been associated with worse educational outcomes in the general population [57]. Additionally, increased educational attainment polygenic risk score demonstrated protective effects across multiple outcomes, including lower total difficulties and reduced conduct problems and hyperactivity at ages 5 and 10 years, as well as better social-emotional development at age 3, consistent with prior evidence linking educational attainment genetic scores with better inhibitory control.[58]

Cleft polygenic score was able to effectively discriminate cases from controls overall but showed heterogeneity across subtypes. While the score performed well for CLO and CLP, it showed less ability to discriminate individuals with cleft palate only from controls. As expected, this reflects the composition of the discovery GWAS meta-analysis used to construct the score, which included CLO and CLP children [11]. The poor performance in CPO is consistent with the established aetiological heterogeneity of orofacial clefts, where CPO is increasingly recognised as genetically distinct from other cleft subtypes [7]. Future studies utilising CPO-specific GWAS would be valuable for developing polygenic scores applicable across all cleft subtypes. Indeed, the results published recently by the Cleft Collective with a CPO-exclusive GWAS [4] should allow future independent studies to demonstrate the validity of such a score. Cleft polygenic risk score also showed little association with behavioural and mental health outcomes within the Cleft Collective, with only modest evidence of association with depressive symptoms at age 10. This finding is consistent with the broader pattern of results suggesting that liability to cleft itself is not linked neurodevelopmental or educational common variant burden [11]

We found little evidence that ND-CNV carriers had differential polygenic risk scores compared to non-carriers for either cognitive, neurodevelopmental traits and psychiatric conditions or cleft. This suggests that rare and common variant effects on cognitive and neuropsychiatric outcomes in cleft operate largely independently. While direct comparisons are limited, studies examining CNV-polygenic score relationships across different phenotypes have similarly found limited interactions; for example, ND-CNVs increase risk of internalising and cardiometabolic multimorbidity largely independently of common variant burden [20]. However, it is worth noting that other studies in clinical cohorts have demonstrated that polygenic scores can provide meaningful risk stratification even in carriers of high-impact rare variants. Studies in ADHD and schizophrenia have found that individuals carrying risk CNVs tend to have lower polygenic scores than cases without these CNVs [59-61]. This is consistent with a liability threshold model; when a high-impact rare variant is present, less common variant burden is required to cross the phenotype threshold. The absence of this pattern in our data may reflect insufficient power given the relatively small number of ND-CNV carriers (n=82). Alternatively, liability threshold dynamics may operate differently here because we are examining neurodevelopmental outcomes as comorbidities within a cleft population, rather than as primary ascertainment phenotypes. In that scenario, both CNV carriers and non-carriers have already crossed the threshold for cleft, and the CNVs additional effect on neurodevelopmental risk may not produce the same polygenic risk score differentiation seen in psychiatric case-control designs.

Mendelian randomization analyses provided no robust evidence that genetic liability to cleft causally influences neurodevelopmental or cognitive outcomes. These findings are in agreement with previous MR analyses on cleft lip with or without cleft palate and educational attainment [11] and demonstrate that it is unlikely that cleft genetic liability is causally linked to neurodevelopmental or cognitive outcomes.

There are several limitations to the present study that should be considered when examining our findings. First, Linkage Disequilibrium Score Regression estimates require large sample sizes for adequate precision [27]. If SNP estimates are imprecise, LDSC can return small and imprecise heritability estimates that preclude reliable estimation of genetic correlations [62]. The Cleft Collective GWAS sample, while substantially large for its field, is modest compared to psychiatric disorder GWAS, potentially limiting its power in detecting weak genetic correlations. However, the consistency of findings across Linkage Disequilibrium Score Regressions, polygenic risk score case-control analyses and MR analyses strengthens confidence in the validity of these results and the conclusion of minimal shared common variant architecture. Second, the behavioural outcome measures rely on parent-reported questionnaires rather than clinical diagnoses which introduces the potential for measurement error. Nevertheless, all instruments utilised are well-validated screening tools with established psychometric properties. Finally, the study population is restricted to European ancestry cases and controls, limiting the generalisability of this study to other populations.

## Conclusions

In summary, this study does not provide evidence for shared common genetic variant architecture between orofacial clefts and cognitive, neurodevelopmental and psychiatric traits. Across multiple complementary approaches—genetic correlation analysis, polygenic risk score comparisons, and Mendelian randomization—we found no support for the hypothesis that shared common genetic variant architecture between orofacial clefts and cognitive, neurodevelopmental and psychiatric traits contributes to the increased neurodevelopmental risk observed in children with CL/P. This stands in contrast to the established enrichment of rare ND-CNVs in this population, particularly among those with cleft palate only. Within the cleft population, polygenic scores for cognitive, neurodevelopmental and psychiatric traits predict behavioural outcomes similarly to the general population, indicating that these genetic factors operate independently of cleft status and are relevant for the cleft population. Given the contribution of rare CNVs to neurodevelopmental comorbidities in cleft, future studies should investigate whether rare single nucleotide variants, particularly de novo coding variants in neurodevelopmental disorder genes, could also be contributing to cognitive and behavioural outcomes in this population.

## Data Availability

Cleft Collective data are available through managed access at https://www.bristol.ac.uk/cleft-collective/professionals/access/. Millennium Cohort Study data are available through the UK Data Service. GWAS summary statistics are publicly available through the Psychiatric Genomics Consortium and other consortia websites. All methods for analyses performed in this manuscript are available in https://github.com/AlexR-genetics/Cleft_commonvariants

## Declarations

### Ethics approval and consent to participate

The Cleft Collective received ethical approval from the South West – Central Bristol NHS Research Ethics Committee (REC reference: 13/SW/0064). Data for the current project were accessed through application CC048-ES. Parents or legal guardians provided written informed consent for participation in the Cleft Collective, including consent for use of biological samples and questionnaire data in approved research projects. The Millennium Cohort Study received ethical approval from the relevant NHS Research Ethics Committees

### Consent for Publication

Not applicable. This study reports aggregate-level statistical analyses only and does not contain individual-level data or identifiable information for any participant.

### Competing Interests

M.J.O. received grants from Akrivia Health, outside the submitted work. All other authors declare that they have no competing interests.

### Funding

This research was funded in whole by the Medical Research Council MRC (MR/W020297/1). For open access purposes, the authors have applied a Creative Commons Attribution (CC BY) public copyright license to any Author Accepted Manuscript (AAM) version arising from this submission. AR, SJL and ES are employed in the MRC Integrative Epidemiology Unit (IEU) (MC_UU_00032/1 - Programme 1: Mendelian Randomization). SJL received support for this study from an MRC project grant (MR/T002093/1). MJO is funded by the Medical Research Council (MR/L010305/1 and programme grant MR/P005748/1). MBMvdB is funded by the Medical Research Council (MR/V004905/1RE). MBMvdB also acknowledges funding from the Tackling Multimorbidity at Scale Strategic Priorities Fund programme (MR/W014416/1) delivered by the Medical Research Council and the National Institute for Health Research in partnership with the Economic and Social Research Council and in collaboration with the Engineering and Physical Sciences Research Council.

### Author’s Contributions

AR, ES and SJL conceived the study. AR performed all statistical analyses (LDSC, polygenic risk score analyses, Mendelian randomization), interpreted the results, and drafted the manuscript. SJL and ES supervised the project, contributed to study design, and critically revised the manuscript. AD, YW, and KH contributed to data collection and phenotypic characterisation within the Cleft Collective. JS contributed to the establishment of the Cleft Collective and critically reviewed the manuscript. GCS and contributed to the genetic data processing and critically reviewed the manuscript. MJO and MBMvdB provided expertise in psychiatric genetics and neurodevelopmental copy number variants and critically revised the manuscript. ES obtained funding and oversaw the study. All authors read and approved the final manuscript.

## Acknowledgements

This publication involves data derived from independent research funded by The Scar Free Foundation; additional funding was provided by The Underwood Trust and the Vocational Training Charitable Trust (VTCT) (REC approval 13/SW/0064). We are grateful to the families who participated in the study, the UK NHS cleft teams, and The Cleft Collective team, who helped facilitate the study. The views expressed in this publication are those of the author(s) and not necessarily those of The Scar Free Foundation, The Underwood Trust, the Vocational Training Charitable Trust or The Cleft Collective Cohort Study team. This work was carried out using the computational facilities of the Advanced Computing Research Centre, University of Bristol - http://www.bristol.ac.uk/acrc/.

## Notes

### Competing Interest Statement

M.J.Owen received grants from Akrivia Health, outside the submitted work. All other authors declare that they have no competing interests.

### Author Declarations

The Cleft Collective received ethical approval from the South West Central Bristol NHS Research Ethics Committee (REC reference: 13/SW/0064). Data for the current project were accessed through application CC048-ES. Parents or legal guardians provided written informed consent for participation in the Cleft Collective, including consent for use of biological samples and questionnaire data in approved research projects. The Millennium Cohort Study received ethical approval from the relevant NHS Research Ethics Committees.

