## Supplemental for "Common Variant Contributions to Neurodevelopmental Risk in Orofacial Clefts"

### Supplemental Material

[Supplemental Table 2. Polygenic Risk Score Regression Models between Cleft PRS (p<0.05 threshold) and all behavioural/developmental outcomes	2](#_Toc225526501)

### Supplemental Table 1: Sample sizes by outcome measure and timepoint

| Outcome | 18 months | 3 years | 5 years | 8 years | 10 years |
| --- | --- | --- | --- | --- | --- |
| SDQ Total | - | - | 571 | 237 | 194 |
| SDQ Conduct | - | - | 573 | 237 | 195 |
| SDQ Emotional | - | - | 573 | 237 | 196 |
| SDQ Hyperactivity | - | - | 571 | 238 | 195 |
| SDQ Peer | - | - | 574 | 239 | 194 |
| SDQ Prosocial | - | - | 574 | 241 | 195 |
| MFQ | - | - | - | - | 196 |
| SCARED | - | - | - | - | 187 |
| ASQ:SE Total | 941 | 706 | 574 | - | - |
| ASQ Communication | 718 | 714 | 576 | - | - |
| ASQ Fine Motor | 715 | 708 | 580 | - | - |
| ASQ Gross Motor | 719 | 712 | 575 | - | - |
| ASQ Personal-Social | 715 | 713 | 579 | - | - |
| ASQ Problem Solving | 700 | 709 | 577 | - | - |

*Note: Sample sizes vary across measures due to questionnaire completion rates at each timepoint.*

### Supplemental Table 2. Polygenic Risk Score Regression Models between Cleft PRS (p<0.05 threshold) and all behavioural/developmental outcomes

| Outcome | Timepoint | β | 95% CI | p-value | N |
| --- | --- | --- | --- | --- | --- |
| SDQ Total | 5 years | -0.10 | -0.66, 0.47 | 0.729 | 571 |
| SDQ Total | 8 years | -0.34 | -1.40, 0.72 | 0.530 | 237 |
| SDQ Total | 10 years | 0.95 | -0.19, 2.08 | 0.101 | 194 |
| SDQ Conduct | 5 years | -0.08 | -0.23, 0.07 | 0.296 | 573 |
| SDQ Conduct | 8 years | -0.04 | -0.30, 0.22 | 0.763 | 237 |
| SDQ Conduct | 10 years | 0.25 | -0.02, 0.53 | 0.075 | 195 |
| SDQ Emotional | 5 years | 0.07 | -0.10, 0.23 | 0.406 | 573 |
| SDQ Emotional | 8 years | -0.03 | -0.38, 0.31 | 0.865 | 237 |
| SDQ Emotional | 10 years | -0.03 | -0.42, 0.36 | 0.880 | 196 |
| SDQ Hyperactivity | 5 years | 0.04 | -0.20, 0.28 | 0.744 | 571 |
| SDQ Hyperactivity | 8 years | -0.08 | -0.53, 0.37 | 0.728 | 238 |
| SDQ Hyperactivity | 10 years | 0.56 | 0.06, 1.06 | 0.028 | 195 |
| SDQ Peer | 5 years | -0.11 | -0.27, 0.05 | 0.178 | 574 |
| SDQ Peer | 8 years | -0.11 | -0.41, 0.19 | 0.472 | 239 |
| SDQ Peer | 10 years | 0.15 | -0.18, 0.49 | 0.380 | 194 |
| SDQ Prosocial | 5 years | 0.20 | 0.01, 0.39 | 0.039 | 574 |
| SDQ Prosocial | 8 years | -0.03 | -0.31, 0.26 | 0.837 | 241 |
| SDQ Prosocial | 10 years | -0.29 | -0.63, 0.06 | 0.099 | 195 |
| MFQ Total | 10 years | 0.69 | 0.06, 1.32 | 0.032 | 196 |
| SCARED Total | 10 years | -0.12 | -2.08, 1.83 | 0.904 | 187 |
| ASQ:SE Total | 18 months | -1.04 | -2.98, 0.90 | 0.293 | 683 |
| ASQ:SE Total | 3 years | -1.62 | -4.80, 1.55 | 0.317 | 706 |
| ASQ:SE Total | 5 years | -2.84 | -7.12, 1.45 | 0.194 | 574 |
| ASQ Communication | 18 months | 0.32 | -0.95, 1.58 | 0.620 | 718 |
| ASQ Communication | 3 years | 0.20 | -0.85, 1.25 | 0.709 | 714 |
| ASQ Communication | 5 years | 0.93 | -0.27, 2.13 | 0.129 | 576 |
| ASQ Fine Motor | 18 months | 0.47 | -0.44, 1.39 | 0.314 | 715 |
| ASQ Fine Motor | 3 years | 0.11 | -1.15, 1.38 | 0.865 | 708 |
| ASQ Fine Motor | 5 years | 1.30 | -0.05, 2.64 | 0.058 | 580 |
| ASQ Gross Motor | 18 months | 0.69 | -0.43, 1.81 | 0.227 | 719 |
| ASQ Gross Motor | 3 years | -0.46 | -1.39, 0.47 | 0.332 | 712 |
| ASQ Gross Motor | 5 years | 1.01 | -0.17, 2.19 | 0.093 | 575 |
| ASQ Personal-Social | 18 months | 0.44 | -0.44, 1.33 | 0.330 | 715 |
| ASQ Personal-Social | 3 years | 0.26 | -0.63, 1.15 | 0.567 | 713 |
| ASQ Personal-Social | 5 years | 0.99 | -0.04, 2.03 | 0.061 | 579 |
| ASQ Problem Solving | 18 months | 0.33 | -0.83, 1.49 | 0.577 | 700 |
| ASQ Problem Solving | 3 years | 0.18 | -0.80, 1.17 | 0.720 | 709 |
| ASQ Problem Solving | 5 years | 0.68 | -0.33, 1.69 | 0.187 | 577 |

*Note: Effect sizes (β) represent change in outcome score per SD increase in Cleft PRS. Models adjusted for sex and 10 principal components.* *For SDQ/MFQ/SCARED and ASQ:SE, higher scores indicate more difficulties; for ASQ domains, higher scores indicate better development.*

### Supplemental Table 3: Association between Polygenic Risk Scores for Cognitive and Neuropsychiatric Traits and Cleft Subtype (Case-Control and Subtype Comparisons)

| Phenotype (1,0) | Variable | Lower_CI | OR | Upper_CI | P_Value |
| --- | --- | --- | --- | --- | --- |
| cleft_lip_only_vs_control | PRS_0.05_ADHD | 0.869 | 0.962 | 1.066 | 0.462 |
| cleft_lip_only_vs_control | PRS_0.05_DEP | 0.951 | 1.048 | 1.156 | 0.345 |
| cleft_lip_only_vs_control | PRS_0.05_ANX | 0.787 | 0.886 | 0.998 | 0.046 |
| cleft_lip_only_vs_control | PRS_0.05_EA | 0.858 | 0.946 | 1.042 | 0.259 |
| cleft_lip_only_vs_control | PRS_0.05_ASD | 0.914 | 1.007 | 1.109 | 0.890 |
| cleft_lip_only_vs_control | PRS_0.05_BP | 0.891 | 0.982 | 1.083 | 0.721 |
| cleft_lip_only_vs_control | PRS_0.05_INT | 0.984 | 1.083 | 1.192 | 0.104 |
| cleft_lip_only_vs_control | PRS_0.05_SCZ | 0.881 | 0.985 | 1.101 | 0.788 |
| cleft_lip_only_vs_cleft_palate_only | PRS_0.05_ADHD | 0.886 | 1.007 | 1.144 | 0.913 |
| cleft_lip_only_vs_cleft_palate_only | PRS_0.05_DEP | 0.975 | 1.100 | 1.241 | 0.123 |
| cleft_lip_only_vs_cleft_palate_only | PRS_0.05_ANX | 0.713 | 0.824 | 0.953 | 0.009 |
| cleft_lip_only_vs_cleft_palate_only | PRS_0.05_EA | 0.884 | 0.998 | 1.126 | 0.971 |
| cleft_lip_only_vs_cleft_palate_only | PRS_0.05_ASD | 0.943 | 1.067 | 1.207 | 0.302 |
| cleft_lip_only_vs_cleft_palate_only | PRS_0.05_BP | 0.919 | 1.038 | 1.173 | 0.550 |
| cleft_lip_only_vs_cleft_palate_only | PRS_0.05_INT | 0.920 | 1.035 | 1.165 | 0.562 |
| cleft_lip_only_vs_cleft_palate_only | PRS_0.05_SCZ | 0.918 | 1.050 | 1.201 | 0.476 |
| cleft_lip_only_vs_cleft_lip_and_palate | PRS_0.05_ADHD | 0.861 | 0.980 | 1.117 | 0.764 |
| cleft_lip_only_vs_cleft_lip_and_palate | PRS_0.05_DEP | 0.850 | 0.959 | 1.081 | 0.490 |
| cleft_lip_only_vs_cleft_lip_and_palate | PRS_0.05_ANX | 0.766 | 0.890 | 1.033 | 0.125 |
| cleft_lip_only_vs_cleft_lip_and_palate | PRS_0.05_EA | 0.868 | 0.978 | 1.100 | 0.706 |
| cleft_lip_only_vs_cleft_lip_and_palate | PRS_0.05_ASD | 0.860 | 0.970 | 1.094 | 0.619 |
| cleft_lip_only_vs_cleft_lip_and_palate | PRS_0.05_BP | 0.873 | 0.986 | 1.114 | 0.821 |
| cleft_lip_only_vs_cleft_lip_and_palate | PRS_0.05_INT | 0.935 | 1.053 | 1.186 | 0.398 |
| cleft_lip_only_vs_cleft_lip_and_palate | PRS_0.05_SCZ | 0.841 | 0.964 | 1.105 | 0.598 |
| cleft_palate_only_vs_control | PRS_0.05_ADHD | 0.885 | 0.960 | 1.041 | 0.319 |
| cleft_palate_only_vs_control | PRS_0.05_DEP | 0.884 | 0.955 | 1.032 | 0.244 |
| cleft_palate_only_vs_control | PRS_0.05_ANX | 0.974 | 1.070 | 1.176 | 0.158 |
| cleft_palate_only_vs_control | PRS_0.05_EA | 0.872 | 0.942 | 1.018 | 0.132 |
| cleft_palate_only_vs_control | PRS_0.05_ASD | 0.876 | 0.947 | 1.023 | 0.166 |
| cleft_palate_only_vs_control | PRS_0.05_BP | 0.875 | 0.946 | 1.022 | 0.157 |
| cleft_palate_only_vs_control | PRS_0.05_INT | 0.968 | 1.045 | 1.128 | 0.258 |
| cleft_palate_only_vs_control | PRS_0.05_SCZ | 0.852 | 0.931 | 1.017 | 0.111 |
| cleft_lip_and_palate_vs_control | PRS_0.05_ADHD | 0.897 | 0.975 | 1.059 | 0.548 |
| cleft_lip_and_palate_vs_control | PRS_0.05_DEP | 1.011 | 1.094 | 1.183 | 0.026 |
| cleft_lip_and_palate_vs_control | PRS_0.05_ANX | 0.894 | 0.985 | 1.084 | 0.755 |
| cleft_lip_and_palate_vs_control | PRS_0.05_EA | 0.895 | 0.968 | 1.047 | 0.417 |
| cleft_lip_and_palate_vs_control | PRS_0.05_ASD | 0.965 | 1.043 | 1.129 | 0.288 |
| cleft_lip_and_palate_vs_control | PRS_0.05_BP | 0.922 | 0.997 | 1.078 | 0.940 |
| cleft_lip_and_palate_vs_control | PRS_0.05_INT | 0.952 | 1.029 | 1.112 | 0.469 |
| cleft_lip_and_palate_vs_control | PRS_0.05_SCZ | 0.925 | 1.013 | 1.108 | 0.785 |

Odds ratios (OR) represent the change in odds of the specified cleft phenotype comparison per standard deviation increase in polygenic risk score (PRS). Models adjusted for sex and ten principal components. Comparisons include each cleft subtype versus population controls, and pairwise comparisons between cleft subtypes. ADHD = Attention-Deficit/Hyperactivity Disorder; ASD = Autism Spectrum Disorder; BP = Bipolar Disorder; DEP = Depression; anx = Anxiety; EA = Educational Attainment; INT = Intelligence; SCZ = Schizophrenia; CI = confidence interval. P-values are uncorrected for multiple testing.

### Supplemental Table 4: Mendelian Randomisation Instrument Strength (F-statistics)

F-statistics for each instrumental variable, calculated as F = β²/SE². All instruments have F > 10, indicating adequate strength to avoid weak instrument bias to conduct MR analysis.

| SNP | CHR | β (exposure) | SE | F-statistic |
| --- | --- | --- | --- | --- |
| rs10172734 | 2 | 0.223 | 0.036 | 39.1 |
| rs17242358 | 8 | 0.405 | 0.041 | 98.8 |
| rs2600519 | 15 | -0.211 | 0.036 | 33.8 |
| rs28361060 | 6 | -0.288 | 0.050 | 33.3 |
| rs3845359 | 1 | 0.199 | 0.034 | 34.7 |
| rs7870540 | 9 | -0.261 | 0.040 | 43.7 |
| rs79482068 | 3 | 0.358 | 0.062 | 33.1 |
| rs9439713 | 1 | 0.239 | 0.035 | 47.0 |

*Mean F-statistic = 45.4; Median = 36.9; Range = 33.1-98.8. All F > 10.*

### Supplemental Table 5: Full Two-Sample Mendelian Randomization Results

| Outcome | N SNPs | IVW | Weighted Median | Weighted Mode | MR-Egger Intercept | MR-Egger Slope† | Heterogeneity |
| --- | --- | --- | --- | --- | --- | --- | --- |
| ADHD | 8 | β=-0.022 (0.014), p=0.120 | β=-0.012 (0.018), p=0.499 | β=-0.002 (0.027), p=0.935 | -0.021 (0.016), p=0.241 | — | Q=3.0, df=7, p=0.886 |
| Autism | 8 | β=0.048 (0.021), p=0.026 | β=0.036 (0.028), p=0.207 | β=0.038 (0.042), p=0.397 | -0.006 (0.024), p=0.798 | — | Q=5.2, df=7, p=0.638 |
| Schizophrenia | 7 | β=0.012 (0.019), p=0.528 | β=0.020 (0.022), p=0.347 | β=0.035 (0.030), p=0.287 | 0.004 (0.022), p=0.852 | — | Q=8.8, df=6, p=0.186 |
| Bipolar Disorder | 8 | β=0.032 (0.021), p=0.122 | β=0.036 (0.017), p=0.034 | β=0.042 (0.022), p=0.094 | -0.022 (0.024), p=0.393 | — | Q=22.0, df=7, p=0.003* |
| Depression | 8 | β=0.004 (0.008), p=0.610 | β=-0.005 (0.007), p=0.483 | β=-0.007 (0.011), p=0.569 | -0.018 (0.006), p=0.019* | β=0.070 (0.021), p=0.017* | Q=16.1, df=7, p=0.024* |
| Anxiety | 8 | β=0.069 (0.038), p=0.069 | β=0.074 (0.046), p=0.110 | β=0.075 (0.061), p=0.258 | 0.0004 (0.042), p=0.992 | — | Q=1.9, df=7, p=0.967 |
| Educational Attainment | 7 | β=0.005 (0.005), p=0.374 | β=-0.001 (0.004), p=0.779 | β=-0.001 (0.005), p=0.914 | 0.006 (0.005), p=0.300 | — | Q=20.9, df=6, p=0.002* |
| Intelligence | 7 | β=-0.002 (0.009), p=0.798 | β=-0.003 (0.007), p=0.683 | β=-0.005 (0.010), p=0.653 | -0.015 (0.008), p=0.124 | — | Q=22.2, df=6, p=0.001* |

*Results presented as β (SE), p-value. β represents the effect of genetic liability to cleft on the log-odds of the outcome. IVW, inverse variance weighted (primary analysis); MR-Egger intercept tests for directional pleiotropy; † MR-Egger slope reported only when the intercept test indicates significant directional pleiotropy (p<0.05). When pleiotropy is absent, the inverse variance weighted estimate is preferred due to greater statistical efficiency. Only depression showed evidence of directional pleiotropy (intercept p=0.019), so the MR-Egger slope is reported for this outcome. Heterogeneity assessed using Cochran's Q statistic. *p<0.05.*

### Supplemental Table S6: Influential Individual SNPs Identified in Leave-One-Out Mendelian Randomisation Analyses

| SNP | Outcome | β | P-value | Notes |
| --- | --- | --- | --- | --- |
| rs10172734 | Educational Attainment | 0.038 | 9.3×10⁻⁶ | Found within the CYFIP Related Rac1 Interactor A (CYRIA) gene 3 Prime UTR region. Variant implicated in multiple studies of Cleft; also reported effect of CYRIA on height, cortical thickness and brain attribute. |
| rs10172734 | Intelligence | -0.053 | 7.7×10⁻⁵ |  |
| rs28361060 | Bipolar Disorder | 0.121 | 1.8×10⁻⁴ | Found in intronic region of TSBP1. TSBP1 implicated in bone tissue density and hypothyroidism |
| rs17242358 | Depression | 0.034 | 7.6×10⁻⁴ | 500B Downstream Variant of LINC00976. LINC00976 is linked to height and monocyte count. |

### Supplemental Figure 1: Mendelian randomization results for causal effects of genetic liability to cleft on neurodevelopmental and psychiatric outcomes

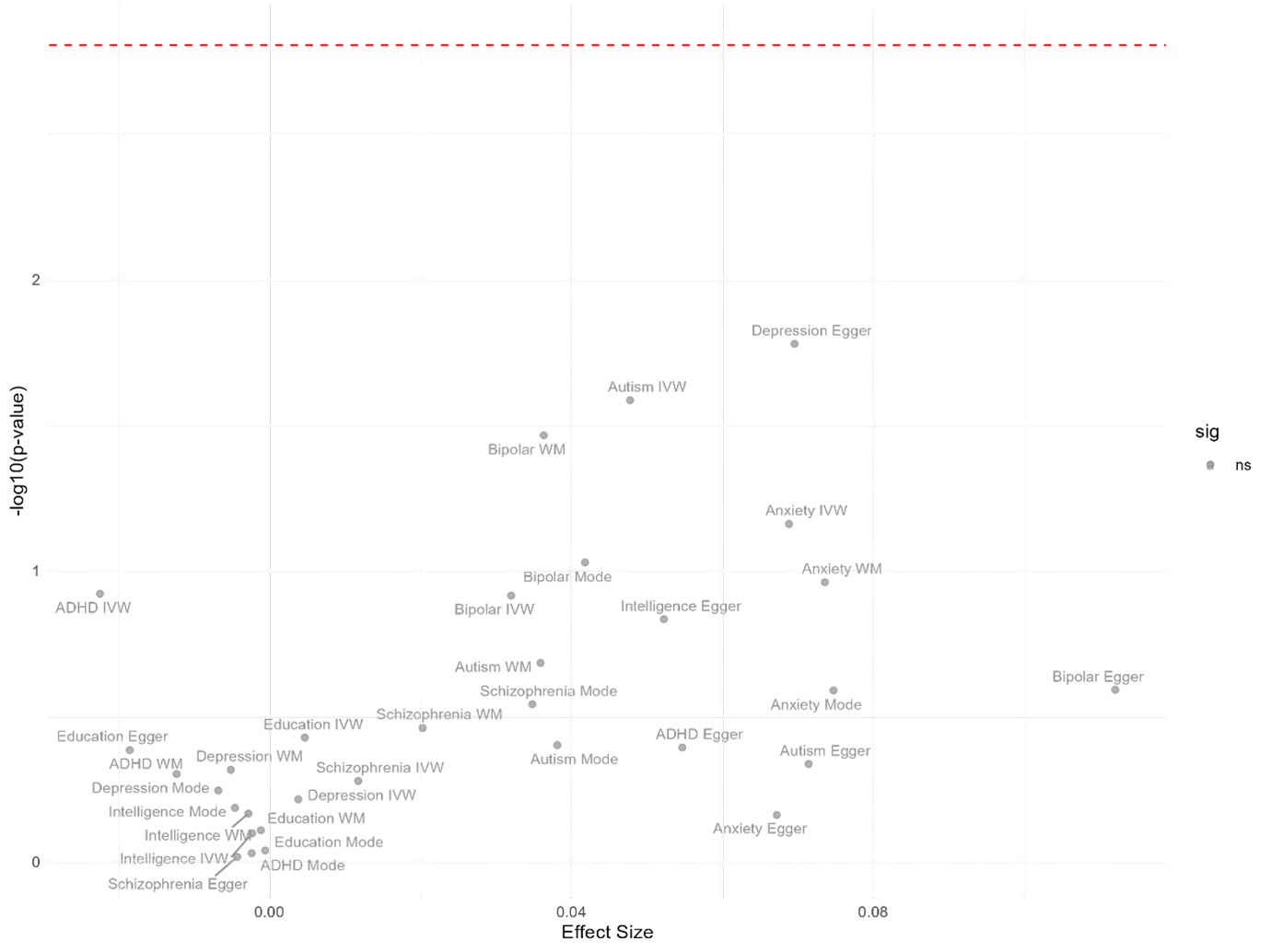

Volcano plot showing effect sizes (beta coefficients, x-axis) and statistical significance (-log10 p-value, y-axis) from two-sample Mendelian randomization analyses testing causal effects of genetic liability to cleft lip/palate on neurodevelopmental and psychiatric outcomes. Results shown for inverse variance weighted (IVW), MR Egger, weighted median (WM), and weighted mode methods. The horizontal red dashed line indicates the Bonferroni correction threshold for multiple testing (p = 0.00156 for 32 comparisons, derived from 8 outcomes × 4 MR methods). Points are colored by statistical significance (grey = non-significant, red = nominally significant p < 0.05). No associations survived correction for multiple testing. Positive effect sizes indicate that genetic liability to cleft increases risk for the outcome. Analysis used eight genome-wide significant cleft variants as instrumental variables.
